# Derivation and validation of a prediction rule for sedative-associated delirium during acute respiratory failure requiring mechanical ventilation

**DOI:** 10.1101/2024.09.30.24314628

**Authors:** Niall T. Prendergast, Chukwudi A. Onyemekwu, Kelly M. Potter, Christopher A. Franz, Georgios D. Kitsios, Bryan J. McVerry, Pratik P. Pandharipande, E. Wesley Ely, Timothy D. Girard

## Abstract

**Background:** Delirium during acute respiratory failure is common and morbid. Pharmacologic sedation is a major risk factor for delirium, but some sedation is often necessary for the provision of safe care of mechanically ventilated patients. A simple, transparent model that predicts sedative-associated delirium in mechanically ventilated ICU patients could be used to guide decisions about personalized sedation.

**Research Question:** Can the risk of sedative-associated delirium be estimated in mechanically-ventilated ICU patients?

**Study Design and Methods:** Using the subset of patients in a previously-published ICU cohort who received mechanical ventilation, we performed backward stepwise logistic regression to derive a model predictive of sedative-associated delirium. We validated this model internally using hundredfold bootstrapping. We then validated this model externally in a separate prospective cohort of mechanically ventilated ICU patients.

**Results:** 836 patients comprised the derivation cohort. Backwards stepwise regression produced a model with age, BMI, sepsis, SOFA, malignancy, COPD, stroke, sex, and doses of sedatives (opioids, propofol, and/or benzodiazepines) as predictors of sedative-associated delirium. The model had very good discriminative power, with an area under the receiver-operator curve (AUROC) of 0.83. Internal validation via bootstrapping showed preserved discriminatory function with an AUROC of 0.81 and graphical evidence of good calibration. External validation in a separate set of 340 patients showed good discrimination, with AUROC of 0.70.

**Interpretation:** Sedative-associated delirium during acute respiratory failure requiring mechanical ventilation can be predicted using a simple, transparent model, which can now be validated in a prospective study.

## Introduction

Delirium, a syndrome of acutely altered attention, awareness, and cognition, affects up to 80% of patients with acute respiratory failure requiring mechanical ventilation.^1-4^ Delirium is associated with both ICU and hospital mortality.^5,6^ Among survivors, delirium is also associated with long-term cognitive impairment,^2,7^ which reduces quality of life and, for many patients,^8^ is a less desirable outcome than death.

During acute respiratory failure, delirium is likely a manifestation of different or converging pathophysiologic processes. Recognizing this multifaceted pathophysiology, clinicians and researchers often subcategorize delirium by probable etiology, with common examples including alcohol withdrawal delirium, septic delirium, delirium associated with hepatic dysfunction (also known as “hepatic encephalopathy”), and medication-induced delirium. In a recent study with a large cohort of ICU patients, 90% of whom were mechanically ventilated, we found that sedative-associated delirium was the most common risk factor-defined delirium subtype.^9^ Like hypoxic and septic delirium, sedative-associated delirium was associated with decreased long-term cognitive function after recovery from critical illness.^9^

In contrast to other etiologies of delirium, sedative-associated delirium is at least in part under the control of the clinical team. However, deliriogenic sedation practices, such as frequent use of benzodiazepines, persist worldwide—indeed, this has worsened since the beginning of the COVID-19 pandemic.^10^ Sedative choice and depth is ideally personalized according to each patient’s condition and needs, but evidence-based tools to guide sedation practices are limited to sedation scales used to gauge depth of sedation.^11,12^ Therefore, we sought to derive and validate a simple, transparent prediction tool that quantifies the risk of sedative-associated delirium during invasive mechanical ventilation, with a particular focus on the newly intubated patient. If accurate and reliable, this tool could then inform clinicians as they consider risk-benefit tradeoffs regarding sedation, particularly in the immediate post-intubation period.

## Methods

### Patient Cohorts

We derived and internally validated the model using data collected during the Bringing to Light the Risk Factors and Incidence of Neuropsychological Dysfunction in intensive care unit (ICU) Survivors (BRAIN-ICU)^2^ and Delirium and Dementia in Veterans Surviving ICU Care (MIND-ICU) studies.^13^ These were parallel multicenter, prospective cohort studies with identical inclusion/exclusion criteria in different patient populations that have been extensively described elsewhere.^2^ In brief, adults with acute respiratory failure and/or septic or cardiogenic shock were enrolled in five US hospitals; complete medication administration data and daily delirium assessments by trained research personnel were collected. BRAIN-ICU and MIND-ICU also collected neurocognitive data out to one year from ICU discharge, as the aims of the study were to estimate the prevalence of long-term cognitive impairment after critical illness and to test the hypothesis that longer duration of delirium in the hospital and higher doses of sedative and analgesic agents are independently associated with more severe cognitive impairment. For the current study, we used only data from ICU admission and restricted the analysis to patients who were mechanically ventilated at the time of study enrollment.

Following derivation and internal validation, we externally validated the rule using data collected as part of the Acute Lung Injury Registry (ALIR),^14^ an ongoing prospective cohort study conducted at the University of Pittsburgh. Inclusion criteria, exclusion criteria, and data gathering procedures have previously been described.^14^ In brief, adults with acute respiratory failure are enrolled from three hospitals within the UPMC health system in Pittsburgh, PA. For the current study, we analyzed mechanically ventilated patients with delirium assessments.

### Baseline Data and Outcomes

During BRAIN-ICU and MIND-ICU, research personnel collected baseline variables at the time of study enrollment, including age, sex, height, weight, admission diagnosis, chronic disease burden according to the Charlson comorbidity index,^15^ preexisting cognitive impairment according to the Short Informant Questionnaire on Cognitive Decline in the Elderly (IQCODE),^16^ and cerebrovascular disease burden according to the Framingham Stroke Risk Profile.^17^ Every study day until ICU discharge or study day 30, research personnel collected data on severity of illness according to the Sequential Organ Failure Assessment (SOFA) score and presence of severe sepsis, hypoxemia, coma, and delirium.^18^ We assessed participants’ level of consciousness with the Richmond Agitation-Sedation Scale (RASS) and assessed for delirium using the Confusion Assessment Method for the ICU (CAM-ICU).^11,19^ We also collected medication-administration data from the medical record, converting benzodiazepine doses to midazolam equivalents and opioid doses to fentanyl equivalents as per the study’s Supplementary Appendix.^2^ As previously described,^9^ we defined sedative-associated delirium as delirium (identified by a positive CAM-ICU assessment) occurring on the same day that the subject received a benzodiazepine, opioid, propofol, and/or dexmedetomidine.

ALIR study personnel collect baseline variables at the time of enrollment, including age, sex, chronic disease burden, height, and weight. Respiratory failure diagnoses are established via a consensus conference of intensivist investigators. Medication data are automatically abstracted from the medical record; doses of benzodiazepines and opioids are converted to midazolam and fentanyl equivalents (respectively) as previously described.^20-22^ For the validation cohort, we included ALIR patients who were assessed for coma and delirium daily using the RASS and CAM-ICU by research personnel and/or twice daily using the Riker Sedation-Agitation Scale (SAS) and the Intensive Care Delirium Screening Checklist (ICDSC).^12,23^ As the ICDSC evaluates a patient over a period of time, while the CAM evaluates a patient at a particular moment, and as delirium is frequently underrecognized, we treated patients as having been delirious on a given day if either CAM-ICU or ICDSC were positive and classified this as sedative-associated delirium using the previously described definition.

## Statistical Analysis

We analyzed data using Stata (version 18 StataCorp, College Station, TX) and R (version 4.1.2, R Foundation for Statistical Computing, Vienna, Austria). Missing BRAIN-ICU data were imputed as previously described.^2^ We imputed missing delirium data in the ALIR dataset (for approximately 9% of measurements) using the MICE package for multivariate imputation in R.

We chose an initial 28 candidate variables that had previously been shown to associate with delirium risk.^24^ Because we sought to derive a relatively simple, transparent score to facilitate implementation, we applied logistic regression so that the resulting model would be intuitively interpretable, the strengths of individual predictors would be explicit, and the model could be used for new prediction in different data sets. Specifically, we used backwards logistic regression with variable elimination at a threshold of p > 0.2 to streamline the model. We used the entire derivation set to create the initial model and used hundred-fold bootstrapping to internally validate the model. Some data (notably frailty and history of stroke) were not captured in the validation cohort, therefore probabilities were calculated in the external validation cohort without those terms. Finally, for both internal and external validation, we assessed discrimination using the area under the receiver-operator curve (i.e., the C-statistic) and assessed calibration graphically and with the Brier index.

### Ethics Approval and Consent to Participate

The BRAIN-ICU and MIND-ICU data comprising the derivation set were obtained with Institutional Review Board (IRB) approval from Vanderbilt University and participating Department of Veterans Affairs Medical Centers. Patients or their legally authorized representatives granted consent, as previously described.^2^ The ALIR study was approved by the IRB at the University of Pittsburgh. Patients or their legally authorized representatives provided initial consent, as previously described.^14^ We also obtained a separate IRB approval from the University of Pittsburgh to analyze deidentified data for the current study.

## Results

Of the 1040 patients in the BRAIN-ICU/MIND-ICU data set, 836 were intubated and mechanically ventilated on the day of enrollment and were used to generate the prediction model. Baseline data are displayed in **Table 1**. Of these 836 patients, 571 (68%) had sedative-associated delirium in the first 72 hours after study enrollment.

**Table 1.** Baseline characteristics and outcome data in generation and validation sets.

| Variable | BRAIN-ICU/MIND-ICU | ALIR |
| --- | --- | --- |
| **Number of patients used** | 836 | 340 |
| **Age, years** | 63.3 [53.7, 72.6] | 59.4 [47.8, 68.4] |
| **Male sex** | 505 (60.4%) | 193 (56.8%) |
| **BMI, kg/m<sup>2</sup>** | 28.9 [24.4, 34.8] | 29.7 [25.6, 36.3] |
| **Day 1 SOFA** | 9 [7, 12] | 7 [4, 10] |
| **History of malignancy** | 180 (21.5%) | 15 (4.4%) |
| **History of COPD** | 254 (30.4%) | 70 (20.6%) |
| **Sepsis at admission** | 159 (31.0%) | 206 (60.6%) |
| **Admission diagnosis** |  |  |
| Acute MI/CHF/arrhythmia | 118 (14.1%) | 35 (10.3%) |
| Airway protection | 108 (12.9%) | 55 (16.2%) |
| ARDS | 301 (36.0%) | 103 (30.3%) |
| Cirrhosis | 15 (1.8%) | 0 (0%) |
| Non-ARDS respiratory | 100 (12.0%) | 147 (43.2%) |
| Other* | 27 (3.2%) | 0 (0%) |
| Surgical | 167 (20.0%) | 0 (0%) |
| **Received benzodiazepines in first 24hrs** | 364 (43.7%) | 288 (84.7%) |
| **Received propofol in first 24hrs** | 360 (43.2%) | 235 (67.6%) |
| **Received opioids in first 24hrs** | 572 (68.6%) | 312 (91.8%) |
| **Sedative-associated delirium in first 72hrs** | 686 (82.1%) | 242 (71.2%) |

Data are displayed as N (%) or median [interquartile range]. Abbreviations: BMI: Body mass index. SOFA: Sequential Organ Failure Assessment. MI: Myocardial infarction. CHF: Congestive heart failure. ARDS: Acute respiratory distress syndrome. COPD: Chronic obstructive pulmonary disease.
\*Other admission diagnoses included hemorrhage, neurological disease, renal failure, malignancy, and other infectious disease.

The 28 initial candidate variables we analyzed in the initial regression are shown in **Table 2**. The strongest predictors were benzodiazepine and propofol doses as well as severity of illness (as measured by SOFA score); other predictors included opioid dose, malignancy, COPD, sepsis at admission, sex, and age (**Table 3**).

**Table 2.**
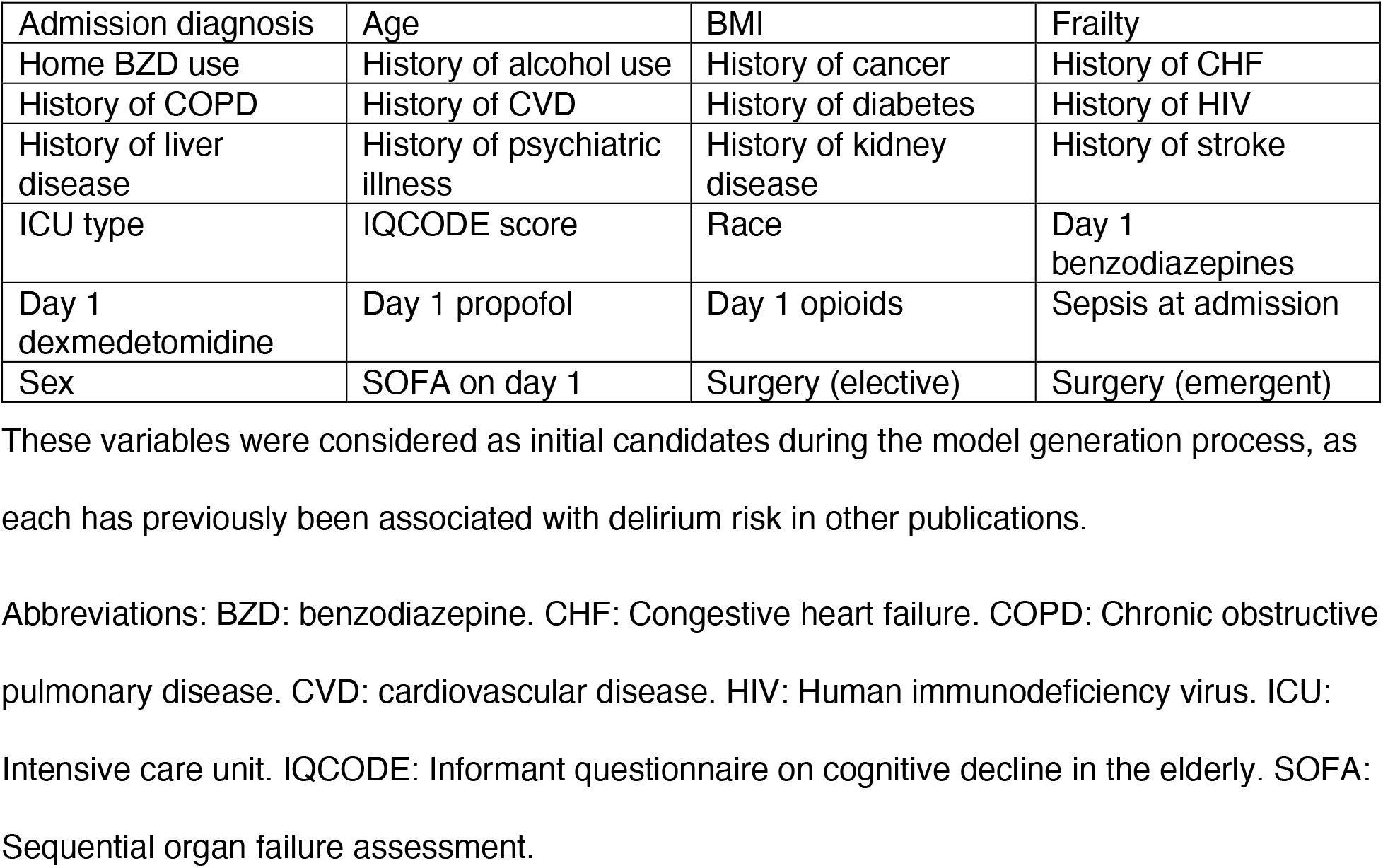
Initial candidate variables prior to backward regression.

|  |  |  |  |
| --- | --- | --- | --- |
| Admission diagnosis | Age | BMI | Frailty |
| Home BZD use | History of alcohol use | History of cancer | History of CHF |
| History of COPD | History of CVD | History of diabetes | History of HIV |
| History of liver disease | History of psychiatric illness | History of kidney disease | History of stroke |
| ICU type | IQCODE score | Race | Day 1 benzodiazepines |
| Day 1 dexmedetomidine | Day 1 propofol | Day 1 opioids | Sepsis at admission |
| Sex | SOFA on day 1 | Surgery (elective) | Surgery (emergent) |

These variables were considered as initial candidates during the model generation process, as each has previously been associated with delirium risk in other publications.
Abbreviations: BZD: benzodiazepine. CHF: Congestive heart failure. COPD: Chronic obstructive pulmonary disease. CVD: cardiovascular disease. HIV: Human immunodeficiency virus. ICU: Intensive care unit. IQCODE: Informant questionnaire on cognitive decline in the elderly. SOFA: Sequential organ failure assessment.

**Table 3.** Variables and associated odds ratios included in the final model.

| Variable | Odds Ratio | 95% Confidence Interval |
| --- | --- | --- |
| Age (per year) | 1.03 | (1.01, 1.04) |
| BMI (per point) | 0.98 | (0.96, 1.00) |
| History of Cancer | 2.22 | (1.24, 3.98) |
| History of COPD | 0.69 | (0.44, 1.09) |
| Sepsis at admission | 1.54 | (0.94, 2.53) |
| Sex (female vs. male) | 1.55 | (1.00, 2.39) |
| SOFA (per point) | 1.19 | (1.11, 1.28) |
| Benzodiazepines<sup>a</sup> | 1.55 | (1.27, 1.88) |
| Opioids<sup>b</sup> | 1.54 | (1.25, 1.90) |
| Propofol<sup>c</sup> | 1.50 | (1.29, 1.73) |

Abbreviations: BMI: Body mass index. COPD: Chronic obstructive pulmonary disease. SOFA:
Sequential organ failure assessment.
<sup>a</sup>Per 10 milligrams of midazolam equivalent received on day 1 of mechanical ventilation
<sup>b</sup>Per milligram of fentanyl equivalent received on day 1 of mechanical ventilation
<sup>c</sup>Per gram of propofol received on day 1 of mechanical ventilation
Note that variables were excluded from the model above a threshold p value of 0.2, therefore
some variables included in the final model have 95% confidence intervals which cross 1.

Concordance statistic analysis showed good discrimination, with an area under the receiver-operator curve of 0.83 (**Figure 1**). Internal validation via hundredfold bootstrapping showed a C-statistic of 0.81 with good graphical calibration (**Figure 2**). External validation in the ALIR cohorts also showed good discrimination, with a C-statistic of 0.70 (**Figure 3**).

**Figure 1.**
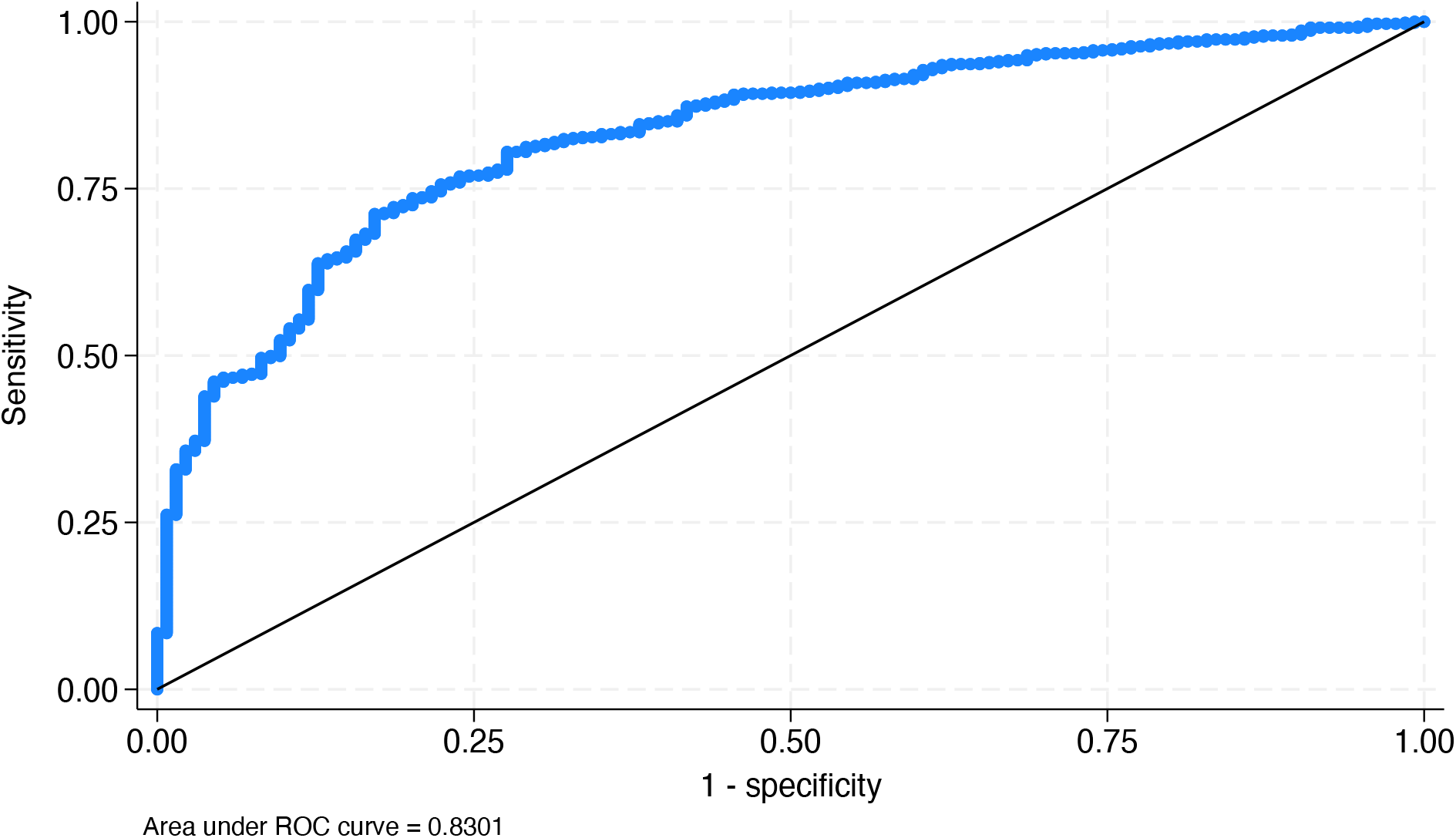
Receiver-operator characteristic curve for training set.

**Figure 2.**
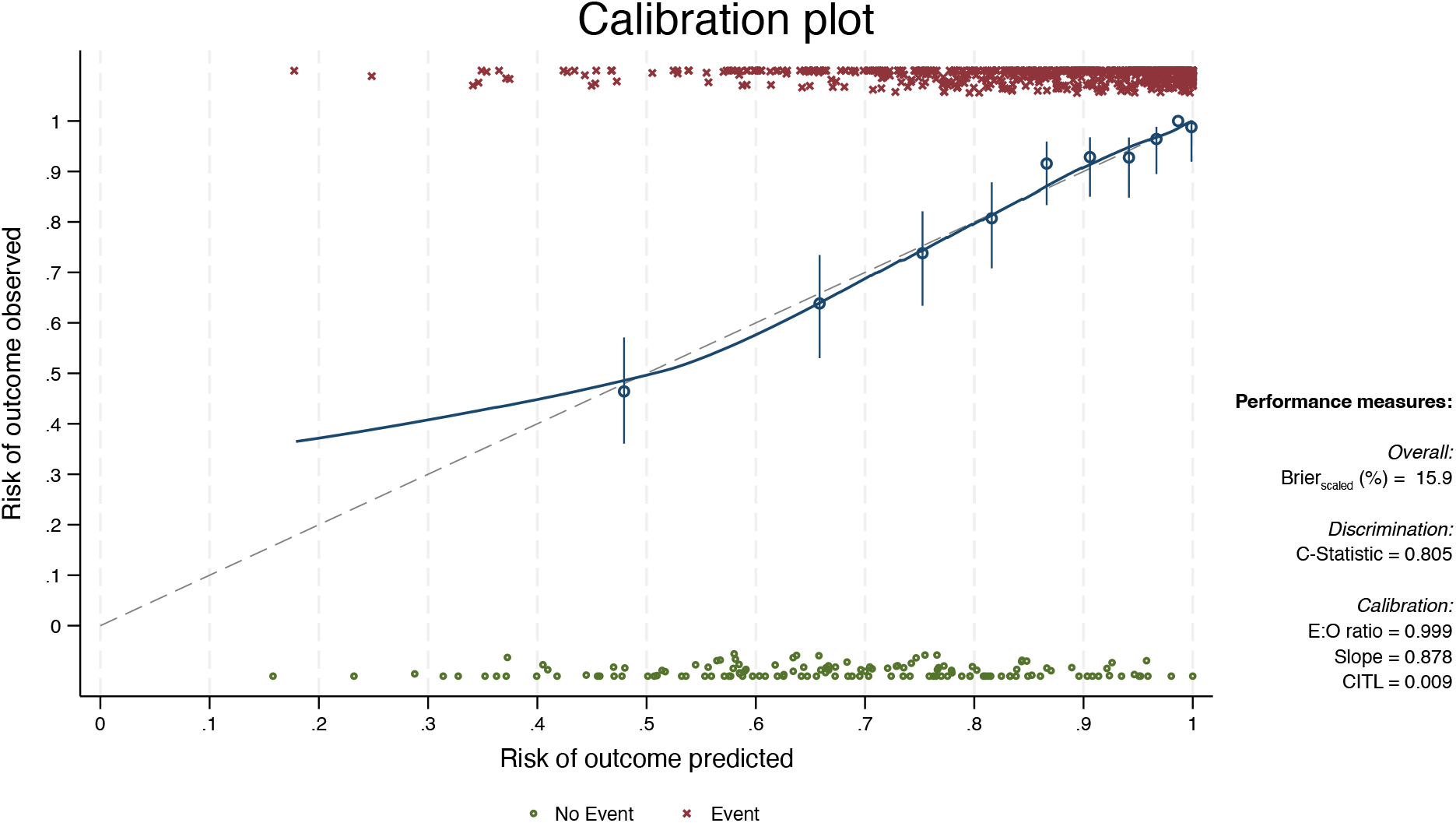
Calibration plot after internal validation using hundredfold bootstrapping.

**Figure 3.**
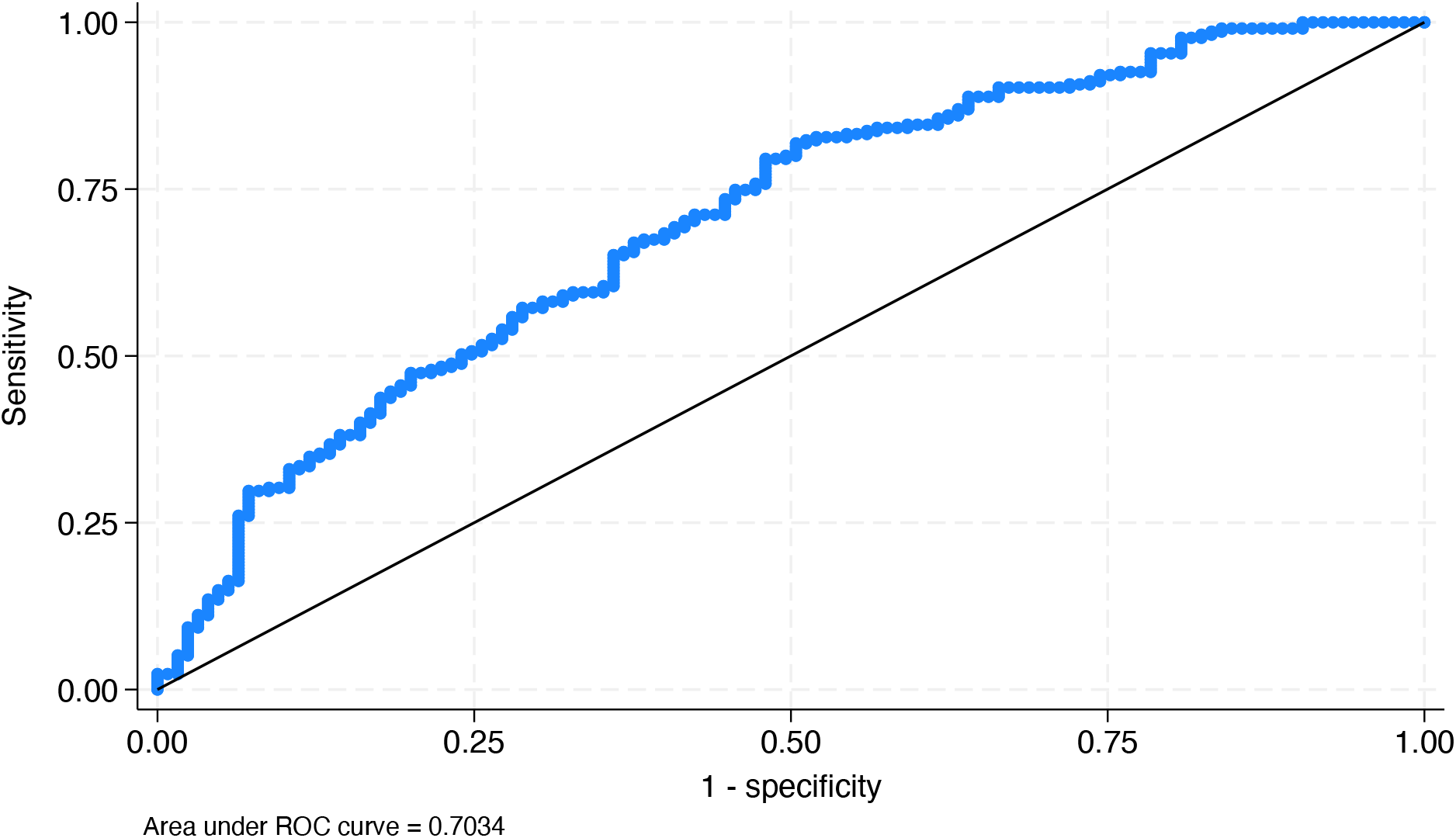
Receiver-operator characteristic curve in external validation data set.

## Discussion

In this study, we found that a 10-variable clinical prediction tool accurately predicted sedative-associated delirium in two multihospital cohorts of mechanically ventilated acute respiratory failure patients. Variables that are readily available to clinicians—age, admission diagnosis, severity of illness (as measured by the SOFA score), malignancy, COPD, sepsis, sex, and doses of benzodiazepines, propofol, and opioids—successfully predicted sedative-associated delirium during the early period of mechanical ventilation. Future studies are now needed to determine whether a personalized sedation protocol based on this model and other relevant data improves outcomes when used clinically.

The most common, and likely most directly modifiable, cause of delirium during acute respiratory failure is pharmacologic sedation. Despite longstanding guidance by multiple professional societies, much of the sedation provided in ICUs around the world is not concordant with guidelines and often places patients at higher risk of delirium.^25^ The reasons for lack of concordance are multiple and complex but may often arise from the need to balance the risks of sedative-associated delirium against other risks inherent in the care of mechanically ventilated patients, for example, the occasional need to maintain deep sedation during management of severe hypoxemic respiratory failure.

Even guideline-concordant sedation is not without delirium risk. Sedation is commonly provided during mechanical ventilation for valid clinical reasons—for example, to treat anxiety or agitation or to avoid excessive work of breathing. Until now, clinicians have not had a way to quantify the delirium risk of any given sedation strategy. Our work allows for the estimation of risk and thus the balancing of that risk against competing interests including work of breathing, metabolic demand, and patient agitation. This tool allows a clinician to estimate the risks for a specific patient associated with multiple different sedation options and thereby to incorporate explicit risk information along with the goals of sedation into his or her decision-making. Thus, this sedation-associated delirium prediction tool could be used to guide decision-making about sedation and may serve as a key element in a personalized sedation protocol, the effects of which should be examined in future clinical trials.

Numerous previous studies have examined risk factors for delirium during critical illness,^26-28^ but none, to our knowledge, have focused specifically on sedative-associated delirium in mechanically ventilated patients. Additionally, though several delirium risk prediction tools have been generated and validated in ICU cohorts,^29-31^ these tools do not distinguish sedative-associated delirium—which is directly related to treatment decisions made by clinicians—from numerous other delirium subtypes and therefore may be less useful as part of a personalized sedation protocol.

Strengths of our study include the use of diverse, high-quality study cohorts for both generation and internal and external validation of the prediction tool. The BRAIN-ICU cohort has been extensively studied and includes data on variables related to patient demographics, comorbidities, acute illness, treatments, and delirium outcomes. The ALIR cohort is a similarly high-quality but fundamentally different cohort, as it is drawn from medical and cardiac intensive care units and includes very few post-surgical patients. The discriminatory power of the model during external validiation thus speaks to its likely generalizability. Other strengths include the tool’s strong face validity and transparency. The predictors included in the final model have all been consistently associated with delirium, and benzodiazepines have been shown to be the highest-risk class of sedative medications used in the intensive care unit, a finding consistent with prior research. One benefit of logistic regression is that it produces clear, transparent effects of each model component. For example, the odds of sedative-associated delirium increase by a factor of 1.55 if ten milligrams of midazolam are used, for example. This transparency should, in theory, make it easier for clinicians to assess the risks of multiple strategies for the same patient and thus to select the lowest-risk strategy that meets the desired sedation goals. Finally, this work is the first to our knowledge to allow for the explicit estimation of sedative-associated delirium risk among mechanically ventilated patients.

One major limitation of this work is the confounding of delirium by coma. Since coma prevents assessment of delirium in the case of a comatose patient, heavy sedation to the point of coma will cause a patient not to score as positive for delirium even if he or she might have been delirious with less sedation. Another limitation of this work include the use of a potentially less predictive statistical approach. More advanced machine learning techniques, such as random forest, extreme gradient boosting, or deep learning neural networks, may fit a given data set better and may generate more accurate predictions. These methods, however, are more prone to overfitting. Additionally, it can be difficult or impossible to interpret the individual variable effects within such models. Along similar lines, the variables available in the two cohorts were not identical; more accurate predictions might be possible with more complete information. A final limitation was inability to include certain other delirium risk factors that were not measured in the cohorts analyzed, such as sensory disturbance or the presence of some invasive devices, such as central venous catheters or fecal collection devices. This limitation could be addressed in prospective validation studies of this model.

## Conclusion

We generated a simple, straightforward prediction tool for early sedative-associated delirium in a diverse cohort of mechanically ventilated patients and validated the tool internally and externally. Our findings suggest that sedative-associated delirium in mechanically ventilated ICU patients can be predicted with good confidence. Since our prediction tool allows treating clinicians to predict the risk of delirium in individual patients for any given sedative strategy, it may allow clinicians to personalize sedation based on explicit risk-benefit tradeoffs. This prediction tool should now be optimized and studied prospectively before its implementation as a clinical decision support tool is tested.

## Data Availability

Availability of data and materials: The datasets used and/or analysed during the current study are available from the corresponding author on reasonable request.

## Declarations

### Ethics approval and consent to participate

BRAIN-ICU and MIND-ICU were approved by the institutional review board (IRB) at Vanderbilt University, and by local IRBs at each site, when the study was initially approved. The use of the data was approved by the IRB at the University of Pittsburgh as PRO18020380. The ALIR was approved as STUDY19050099 by the IRB at the University of Pittsburgh. Both studies involve(d) consent by the patient or his/her legally authorized representative.

### Availability of data and materials

The datasets used and/or analysed during the current study are available from the corresponding author on reasonable request.

### Competing interests

NTP, CAO, KMP, and CAF declare that they have no competing interests. GDK has received research funding from Genentech, Inc, and Pfizer, Inc, unrelated to this work. BJM receives research funding from Genentech, and previously served as consultant or advisory board member for BioAegis, Synairgen Research Ltd, and Boehringer Ingelheim. PPP declares no competing interest. EWE reports honoraria for continuing medical education lectures sponsored by Pfizer, and study support (investigational drug provision, no direct payments) from Eli Lilly. TDG receives research funding from Ceribell and served previously on an advisory board for Lungpacer Medical Inc.

### Funding

This work was supported by NIH grants F32HL158058 (to NTP), R03HL162655 (to GDK), P01HL114453 (to BJM), and R01AG027472 (to EWE) and by a Merit Review Grant from Department of Veterans Affairs (to EWE).

### Authors’ contributions

NTP and TDG conceived of and designed the work. NTP, CAF, GDK, BJM, PPP, EWE, and TDG participated in the acquisition and analysis of data. NTP, CAO, KMP, CAF, GDK, BJM, and TDG participated in the drafting and/or substantial revision of the manuscript. All authors have approved the submitted manuscript.

## Acknowledgements

Not applicable.

## Notes

### Author Declarations

Ethics approval and consent to participate: BRAIN-ICU and MIND-ICU were approved by the institutional review board (IRB) at Vanderbilt University, and by local IRBs at each site, when the study was initially approved. The use of the data was approved by the IRB at the University of Pittsburgh as PRO18020380. The ALIR was approved as STUDY19050099 by the IRB at the University of Pittsburgh. Both studies involve(d) consent by the patient or his/her legally authorized representative.

## References

1. Khan BA, Fadel WF, Tricker JL et al. Effectiveness of implementing a wake up and breathe program on sedation and delirium in the ICU. Crit Care Med. 2014;42(12):e791–795.

2. Pandharipande PP, Girard TD, Jackson JC et al. Long-term cognitive impairment after critical illness. N Engl J Med. 2013;369(14):1306–1316.

3. American Psychiatric Association., American Psychiatric Association. DSM-5 Task Force. Diagnostic and statistical manual of mental disorders : DSM-5. 5th ed. Washington, D.C.: American Psychiatric Association; 2013

4. Prendergast NT, Franz CA, Schaefer C et al. Inflammatory Subphenotype Is Associated with Acute Brain Dysfunction in Mechanically Ventilated Patients. Ann Am Thorac Soc. 2024;21(9):1329–1333.

5. Salluh JI, Wang H, Schneider EB et al. Outcome of delirium in critically ill patients: systematic review and meta-analysis. BMJ. 2015;350:h2538.

6. Ely EW, Shintani A, Truman B et al. Delirium as a predictor of mortality in mechanically ventilated patients in the intensive care unit. JAMA. 2004;291(14):1753–1762.

7. Girard TD, Jackson JC, Pandharipande PP et al. Delirium as a predictor of long-term cognitive impairment in survivors of critical illness. Crit Care Med. 2010;38(7):1513–1520.

8. Fried TR, Bradley EH, Towle VR et al. Understanding the treatment preferences of seriously ill patients. N Engl J Med. 2002;346(14):1061–1066.

9. Girard TD, Thompson JL, Pandharipande PP et al. Clinical phenotypes of delirium during critical illness and severity of subsequent long-term cognitive impairment: a prospective cohort study. Lancet Respir Med. 2018;6(3):213–222.

10. Luz M, Brandao Barreto B, de Castro REV et al. Practices in sedation, analgesia, mobilization, delirium, and sleep deprivation in adult intensive care units (SAMDS-ICU): an international survey before and during the COVID-19 pandemic. Ann Intensive Care. 2022;12(1):9.

11. Sessler CN, Gosnell MS, Grap MJ et al. The Richmond Agitation-Sedation Scale: validity and reliability in adult intensive care unit patients. Am J Respir Crit Care Med. 2002;166(10):1338–1344.

12. Riker RR, Picard JT, Fraser GL. Prospective evaluation of the Sedation-Agitation Scale for adult critically ill patients. Crit Care Med. 1999;27(7):1325–1329.

13. Hughes CG, Patel MB, Jackson JC et al. Surgery and Anesthesia Exposure Is Not a Risk Factor for Cognitive Impairment After Major Noncardiac Surgery and Critical Illness. Ann Surg. 2017;265(6):1126–1133.

14. Kitsios GD, Yang L, Manatakis DV et al. Host-Response Subphenotypes Offer Prognostic Enrichment in Patients With or at Risk for Acute Respiratory Distress Syndrome. Crit Care Med. 2019;47(12):1724–1734.

15. Charlson ME, Pompei P, Ales KL et al. A new method of classifying prognostic comorbidity in longitudinal studies: development and validation. J Chronic Dis. 1987;40(5):373–383.

16. Jorm AF, Scott R, Cullen JS et al. Performance of the Informant Questionnaire on Cognitive Decline in the Elderly (IQCODE) as a screening test for dementia. Psychol Med. 1991;21(3):785–790.

17. Llewellyn DJ, Lang IA, Xie J et al. Framingham Stroke Risk Profile and poor cognitive function: a population-based study. BMC Neurol. 2008;8:12.

18. Vincent JL, Moreno R, Takala J et al. The SOFA (Sepsis-related Organ Failure Assessment) score to describe organ dysfunction/failure. On behalf of the Working Group on Sepsis-Related Problems of the European Society of Intensive Care Medicine. Intensive Care Med. 1996;22(7):707–710.

19. Ely EW, Inouye SK, Bernard GR et al. Delirium in mechanically ventilated patients: validity and reliability of the confusion assessment method for the intensive care unit (CAM-ICU). JAMA. 2001;286(21):2703–2710.

20. Patanwala AE, Duby J, Waters D et al. Opioid conversions in acute care. Ann Pharmacother. 2007;41(2):255–266.

21. Barr J, Zomorodi K, Bertaccini EJ et al. A double-blind, randomized comparison of i.v. lorazepam versus midazolam for sedation of ICU patients via a pharmacologic model. Anesthesiology. 2001;95(2):286–298.

22. Ng K, Dahri K, Chow I et al. Evaluation of an alcohol withdrawal protocol and a preprinted order set at a tertiary care hospital. Can J Hosp Pharm. 2011;64(6):436–445.

23. Bergeron N, Dubois MJ, Dumont M et al. Intensive Care Delirium Screening Checklist: evaluation of a new screening tool. Intensive Care Med. 2001;27(5):859–864.

24. Wilson JE, Mart MF, Cunningham C et al. Delirium. Nat Rev Dis Primers. 2020;6(1):90.

25. Devlin JW, Skrobik Y, Gelinas C et al. Clinical Practice Guidelines for the Prevention and Management of Pain, Agitation/Sedation, Delirium, Immobility, and Sleep Disruption in Adult Patients in the ICU. Crit Care Med. 2018;46(9):e825–e873.

26. Davis DHJ, Skelly DT, Murray C et al. Worsening cognitive impairment and neurodegenerative pathology progressively increase risk for delirium. Am J Geriatr Psychiatry. 2015;23(4):403–415.

27. Persico I, Cesari M, Morandi A et al. Frailty and Delirium in Older Adults: A Systematic Review and Meta-Analysis of the Literature. J Am Geriatr Soc. 2018;66(10):2022–2030.

28. Van Rompaey B, Elseviers MM, Schuurmans MJ et al. Risk factors for delirium in intensive care patients: a prospective cohort study. Crit Care. 009;13(3):R77.

29. Wassenaar A, van den Boogaard M, van Achterberg T et al. Multinational development and validation of an early prediction model for delirium in ICU patients. Intensive Care Med. 2015;41(6):1048–1056.

30. van den Boogaard M, Pickkers P, Slooter AJ et al. Development and validation of PRE-DELIRIC (PREdiction of DELIRium in ICu patients) delirium prediction model for intensive care patients: observational multicentre study. BMJ. 2012;344:e420.

31. Marra A, Pandharipande PP, Shotwell MS et al. Acute Brain Dysfunction: Development and Validation of a Daily Prediction Model. Chest. 2018;154(2):293–301.

